# Community identity, poverty, and antimicrobial discontinuation in a Particularly Vulnerable Tribal Group in central India: a cross-sectional study of healthcare choice and differentiated stewardship

**DOI:** 10.64898/2026.06.24.26356485

**Authors:** D Konar, G Patil, I Pradhan, D Singh, T Singh, B Chatterjee, K Walia, R Nandy, R Kataria

**Affiliations:** Jan Swasthya Sahyog, Ganiyari, India; Indian Council of Medical Research, Division of Epidemiology and Communicable Diseases, New Delhi, India; IQ City Medical College, Department of Microbiology, Durgapur, India; University of North Texas Health Science Center, Department of Biostatistics and Epidemiology, Fort Worth, USA

## Abstract

**Background:** Antimicrobial stewardship in many settings assumes that community antimicrobial misuse reflects low awareness, and favours education-based interventions. Population-level evidence on healthcare seeking and antimicrobial practices in marginalised indigenous groups in low-income and middle-income countries is limited. We examined socioeconomic and identity-related determinants of healthcare-provider choice, antimicrobial awareness, and harmful antimicrobial practices in a tribal population in central India.

**Methods:** We did a cross-sectional survey of 1146 adults in the catchment area of Jan Swasthya Sahyog (JSS), a non-profit community health organisation, in Bilaspur and Mungeli districts, Chhattisgarh, India (January, 2021-April, 2022). Healthcare-provider choice was modelled with binary and multinomial logistic regression (government as reference), and antimicrobial awareness and ten harmful practices with logistic regression, applying Benjamini-Hochberg false discovery rate (FDR) correction within each family. A 30-day treatment-recall sub-study (n=284) assessed actual treatment location and out-of-pocket cost. Models with rare events or separation were refitted with Firth penalised regression.

**Findings:** Median per capita income was INR 8000 per year. Baiga identity was associated with higher odds of using informal (odds ratio 2.58) and private (2.55) providers rather than government facilities, but not JSS (1.36). Awareness of antimicrobials was 7.6% and was associated mainly with education (primary-or-less vs college 0.04). Baiga identity was independently associated with premature discontinuation (4.77), stopping for perceived intolerance (4.43), and financial discontinuation (6.81, 95% CI 3.51-13.25), but with lower odds of stopping because of perceived recovery (0.12). In the sub-study, predicted government-facility use was 1.6% for Baiga versus 18.1% for non-Baiga individuals. Findings were robust to Firth penalisation.

**Interpretation:** In this population, antibiotic non-completion was associated with poverty and access constraints rather than only with awareness, and a non-profit provider appeared to reach groups more equitably. Affordability-oriented, differentiated stewardship merits prospective evaluation. Findings are from a single catchment, are associational, and should be interpreted with the study’s sampling in mind.

**Funding:** Indian Council of Medical Research.

Research in Context

Evidence before this study
We searched PubMed, Embase, and Google Scholar for studies published up to January, 2026, using terms including “antimicrobial use”, “healthcare seeking”, “tribal” or “indigenous”, “Particularly Vulnerable Tribal Group”, and “India”, without language restriction. Population-level quantitative evidence on healthcare-provider choice and antimicrobial practices in marginalised indigenous communities in low-income and middle-income countries is scarce; most available data are from qualitative studies or administrative records and rarely separate the effects of poverty, education, and community identity, or compare public, private, informal, and non-profit providers within the same population.

Added value of this study
In a cross-sectional survey of 1146 adults in a tribal catchment area in central India, Baiga identity (a Particularly Vulnerable Tribal Group) was independently associated with use of informal and private rather than government providers, and with poverty-related, apparently involuntary antibiotic discontinuation, but not with reduced use of a local non-profit provider. Awareness of antimicrobials was very low and was associated chiefly with formal education. These patterns suggest that antibiotic non-completion in this setting is associated with affordability and access constraints rather than only with knowledge.

Implications of all the available evidence
In comparable settings, stewardship approaches focused only on awareness may be insufficient where non-completion reflects cost and access. Community-based non-profit providers may reach marginalised groups more equitably, although this requires confirmation in studies not anchored to a single provider’s catchment. Affordability-oriented, differentiated stewardship - including support for course completion - warrants prospective evaluation.

## Introduction

Antimicrobial resistance is a major threat to health, and its burden falls disproportionately on low-income and middle-income countries (LMICs). Much international stewardship guidance emphasises demand-side measures - improving public awareness, restricting over-the-counter access, and promoting adherence - reflecting an assumption that community antimicrobial misuse is primarily a behavioural problem rooted in limited education.^1.2^ This framing may underweight how poverty, access, and institutional exclusion shape, and sometimes constrain, medicine use.^3,4,5^

India’s Scheduled Tribes comprise about 8.6% of the population but are over-represented in the lowest strata of income, education, and health.^6^ Among them, Particularly Vulnerable Tribal Groups (PVTGs) - such as the Baiga of central India - experience intersecting deprivations, including subsistence economies and limited reach of state institutions. Robust population-level data on healthcare seeking, health literacy, and antimicrobial practices in PVTG communities are scarce; most evidence is qualitative or administrative.

Jan Swasthya Sahyog (JSS) is a non-profit community health organisation in Bilaspur district, Chhattisgarh, providing low-cost primary and secondary care through community health workers. Its catchment area allows examination of how community identity, income, and provider type (government, private, informal, and non-profit) relate to health behaviours. We analysed a cross-sectional survey in this catchment to characterise socioeconomic profile, healthcare-provider choice, antimicrobial awareness, and harmful antimicrobial practices, and to compare the reach of government and non-profit providers among Baiga and non-Baiga residents.

## Methods

### Study design and setting

We did a cross-sectional survey in the catchment area of Jan Swasthya Sahyog (JSS), a non-profit community health organisation whose service area covers rural Bilaspur and Mungeli districts of Chhattisgarh, India, between January, 2021, and April, 2022. Trained field investigators administered a structured questionnaire, translated into local languages, in face-to-face interviews. Reporting follows the STROBE guideline.

### Participants

Participants were recruited on a catchment basis from the population served by JSS. Eligible participants were consenting adult residents of the catchment area - household heads or primary adult caregivers able to provide household income, expenditure, demographic, and health information - with one respondent interviewed per household. Recruitment was conducted through JSS’s community health network across its service villages; respondents originated from 265 villages, predominantly in Bilaspur and Mungeli districts, with a small number from neighbouring districts. This was a non-probability, catchment-based sample rather than a probability sample. Of 1175 individuals approached, 1146 were enrolled (29 declined or were unavailable; 2.5% non-response) and one record with completely missing data was excluded, leaving 1145 for analysis (appendix figure 1). The questionnaire was adapted into the local language and pre-tested with community members in a pilot phase.

### Sample size

The achieved sample of 1146 provides approximately +/-2.5% absolute precision for a population proportion of 20% at 95% confidence under simple-random-sampling assumptions, and satisfies the rule of at least ten events per variable for all multivariable models except the rarest antimicrobial-discontinuation outcomes, which were additionally examined with Firth penalised regression.

### Variables

Per capita household income was calculated as total annual household income divided by household size. Because the household income distribution was highly right-skewed (skewness 8.1), income was summarised using medians, IQRs, and the median absolute deviation. Stated primary healthcare contact was classified into four mutually exclusive categories - government, JSS, informal practitioner, and private clinic; mixed-provider contacts (n=175; 16.5%) and missing responses (n=4) were excluded from the provider-choice models. We assessed five antimicrobial-literacy outcomes and ten harmful antimicrobial-practice outcomes. For branching questions on reasons for non-completion, “not applicable” responses were treated as missing rather than “no”.

### Statistical analysis

Healthcare-provider choice was modelled by binary logistic regression (government vs non-government, with backward elimination at p>0.10) and multinomial logistic regression (government as reference). Awareness and antimicrobial-practice outcomes were modelled separately by logistic regression. Predictors were community identity (Baiga vs non-Baiga), per capita income, education (four categories, college or above as reference), sex, age, chronic disease, and, where relevant, distance to the nearest facility. “Don’t know” responses were excluded outcome by outcome and models used complete cases, so the analytic sample varied by variable; the sociodemographic predictors had no missing values. Odds ratios are presented with 95% CIs and discrimination as the area under the receiver operating characteristic curve (AUC). Adjusted estimates are reported; unadjusted estimates were consistent in direction and are available on request. To limit false-positive findings from multiple testing, we applied Benjamini-Hochberg FDR correction within each analytic family (FDR-adjusted p<0.05 significant). Predicted probabilities were computed at mean covariate values. Analyses used Python 3.

### 30-day treatment-recall sub-study

In an embedded sub-study, respondents reporting an illness episode in the preceding 30 days (n=284) were asked where they sought treatment and what they spent. Actual treatment location was modelled by multinomial regression (single-provider episodes, n=280; government as reference). Usual monthly health expenditure (Q) was compared with out-of-pocket cost for the recalled episode (Z) using the paired Wilcoxon signed-rank test; costs are interpreted descriptively because episode expenditure is endogenous to provider choice.

### Sensitivity analysis

Several models involved rare outcomes or sparse covariate cells, including quasi-complete separation, where standard maximum-likelihood logistic regression can be unstable. As a sensitivity analysis, the branching antimicrobial-discontinuation outcomes and the recall sub-study contact model were refitted with Firth penalised-likelihood regression (appendix table A1).

### Ethics

The study was approved by the Shaheed Hospital institutional ethics committee. Participants gave free and informed consent before enrolment. Community members and community health workers contributed to local-language adaptation and field-testing of the questionnaire and to community sensitisation and recruitment; survey participants were not formally involved in the research question, design, conduct, or analysis.

## Results

### Population and economic profile

Of 1146 individuals, 742 (64.7%) were female, 278 (24.3%) were Baiga, and 746 (65.1%) were of Scheduled Tribe caste; 880 (76.8%) lived in nuclear families (table 1). Educational deprivation was marked: 438 (38.2%) had no formal schooling and only 48 (4.2%) had college or higher qualifications; no formal schooling was more common among Baiga than non-Baiga respondents (55.0% vs 32.3%). Median income and its dispersion increased with educational attainment (table 1).

**Table 1:** Baseline sociodemographic and economic characteristics (N=1146)

**(a) Sociodemographic characteristics**
| Characteristic | Category | n | % |
| --- | --- | --- | --- |
| Sex | Female | 742 | 64.7 |
|  | Male | 404 | 35.3 |
| Age (years) | 17-25 | 208 | 18.2 |
|  | 26-40 | 483 | 42.2 |
|  | 41-55 | 282 | 24.6 |
|  | 56-70 | 138 | 12.0 |
|  | >70 | 35 | 3.1 |
| Community identity | Non-Baiga | 868 | 75.7 |
|  | Baiga | 278 | 24.3 |
| Caste | Scheduled Tribe | 746 | 65.1 |
|  | Other Backward Class | 264 | 23.0 |
|  | Scheduled Caste | 124 | 10.8 |
|  | General | 12 | 1.0 |
| Education | No formal schooling | 438 | 38.2 |
|  | Less than primary | 354 | 30.9 |
|  | High school | 238 | 20.8 |
|  | Secondary | 68 | 5.9 |
|  | College or above | 48 | 4.2 |
| Family type | Nuclear | 880 | 76.8 |
|  | Joint | 210 | 18.3 |
|  | Single/extended | 55 | 4.8 |

**(b) Per capita income by community identity**
| Group | n | Median per capita income (INR/year) | Median household income (INR/year) | Mean household size |
| --- | --- | --- | --- | --- |
| All participants | 1146 | 8000 (IQR 5000-13 333) | 36 000 | 5.17 |
| Non-Baiga | 868 | 8333 | 36 000 | 5.39 |
| Baiga | 278 | 7350 | 30 000 | 4.49 |

*Per capita income = household income / household size; median absolute deviation for all participants was 3917. Income detail by caste, occupation and education is in appendix tables S1-S2.*

Household income was highly right-skewed (median INR 36 000 per year; mean INR 57 668). Median per capita income was INR 8000 per year (IQR 5000-13 333; about INR 667 per month), with 49.1% below this value. Median annual household health expenditure (INR 1200) was 3.6% of median household income but 15% of median per capita income. Baiga households had a lower median per capita income than non-Baiga households (INR 7350 vs INR 8333), a difference that persisted after accounting for household size (table 1).

### Healthcare-provider choice

Among 1040 respondents with a classifiable primary contact, 207 (19.9%) used a government facility and 833 (80.1%) a non-government provider. In the binary model, Baiga identity was the only FDR-significant predictor of non-government contact (odds ratio 1.96, 95% CI 1.28-2.98); income, education, sex, age, and distance were not significant. Backward elimination retained Baiga identity alone (2.11, 1.42-3.16; table 2).

**Table 2:** Healthcare-provider choice.

**(a) Multinomial logistic regression of four-way contact (reference: government)**
| Predictor | OR (95% CI) | p (FDR) | Sig |
| --- | --- | --- | --- |
| **JSS vs government** |  |  |  |
| Baiga (ref non-Baiga) | 1.36 (0.85-2.19) | 0.493 |  |
| Distance to facility (per km) | 1.03 (1.01-1.04) | 0.019 | * |
| Male (ref female) | 1.24 (0.85-1.82) | 0.570 |  |
| Age (per year) | 0.98 (0.97-1.00) | 0.157 |  |
| **Informal practitioner vs government** |  |  |  |
| Baiga (ref non-Baiga) | 2.58 (1.55-4.30) | 0.007 | ** |
| Male (ref female) | 1.73 (1.14-2.64) | 0.064 | . |
| Distance to facility (per km) | 1.00 (0.98-1.02) | 0.985 |  |
| **Private clinic vs government** |  |  |  |
| Baiga (ref non-Baiga) | 2.55 (1.40-4.65) | 0.019 | * |
| Education: secondary (ref college+) | 0.17 (0.03-0.96) | 0.178 |  |
| Education: primary or less (ref college+) | 0.34 (0.09-1.24) | 0.347 |  |

**(b) Predicted probabilities of contact type at mean covariate values (%)**
| Group | Government | JSS | Informal | Private |
| --- | --- | --- | --- | --- |
| Non-Baiga (ref) | 26.3 | 43.9 | 19.9 | 9.9 |
| Baiga | 16.2 | 36.7 | 31.6 | 15.5 |
| College or above (ref) | 17.8 | 31.0 | 27.6 | 23.6 |
| Primary or less | 23.7 | 44.2 | 21.4 | 10.6 |

Probabilities computed at sample-mean covariate values, varying one predictor at a time; rows sum to 100% excluding rounding.

The multinomial model (n=866; government reference) clarified the pattern (table 2). Relative to government facilities, Baiga individuals had higher odds of using informal practitioners (2.58, 1.55-4.30) and private clinics (2.55, 1.40-4.65), whereas JSS use relative to government did not differ by identity (1.36, 0.85-2.19). Greater distance predicted JSS rather than government use (1.03 per km, 1.01-1.04). At mean covariate values, predicted government use was 16.2% among Baiga versus 26.3% among non-Baiga individuals, with the difference distributed across informal (31.6% vs 19.9%) and private (15.5% vs 9.9%) care; JSS remained the leading provider for both groups.

### Antimicrobial awareness and harmful practices

Awareness of antimicrobials was low: 7.6% had heard of antibiotics or antimicrobials. Awareness was associated chiefly with education (primary-or-less vs college 0.04, 0.02-0.09; high school 0.12), and, independently, with Baiga identity (0.17, 0.05-0.54; AUC 0.788; table 3).

**Table 3:** Antimicrobial awareness and harmful practices.

**(a) Healthcare-literacy outcomes: model summary and FDR-significant predictors**
| Outcome (1=desirable) | n | Event % | AUC | FDR-significant predictors (OR) |
| --- | --- | --- | --- | --- |
| Ever heard of antimicrobials | 1131 | 7.6 | 0.788 | Edu primary/less 0.04***; high school 0.12***; secondary 0.30*; Baiga 0.17* |
| Cold: appropriate vs does not know medicine | 1074 | 59.1 | 0.619 | Age 0.98/yr**; per capita income (+)* |
| Diarrhoea: appropriate vs unknowingly takes antibiotics | 626 | 46.5 | 0.609 | Male 0.60* |
| Diarrhoea: appropriate vs no treatment | 761 | 38.2 | 0.595 | None |
| Recalls prescription from last visit | 1016 | 19.9 | 0.575 | None |

(b) FDR-significant associations for harmful antimicrobial practices
| Outcome | Predictor | OR (95% CI) | p (FDR) |
| --- | --- | --- | --- |
| Livestock/agriculture use | Chronic disease | 2.02 (1.32-3.09) | 0.008 |
| Sharing medications | Age (per year) | 0.99 (0.98-1.00) | 0.034 |
| Leftover medicine use | Baiga | 1.93 (1.42-2.64) | <0.001 |
| Leftover medicine use | Age (per year) | 0.98 (0.97-0.99) | 0.008 |
| Premature discontinuation | Baiga | 4.77 (3.39-6.70) | <0.001 |
| Stopping (intolerance) | Baiga | 4.43 (2.50-7.83) | <0.001 |
| Discontinuation (financial) | Baiga | 6.81 (3.51-13.25) | <0.001 |
| Stopping (disease resolved) | Baiga | 0.12 (0.05-0.32) | <0.001 |
| Stopping (disease resolved) | Edu primary or less | 0.09 (0.03-0.24) | <0.001 |
| Stopping (disease resolved) | Edu high school | 0.15 (0.06-0.42) | 0.002 |
| Stopping (disease resolved) | Chronic disease | 3.29 (1.75-6.17) | 0.002 |

FDR across 80 tests (10 models x 8 predictors). Lower per capita income was additionally significant for leftover-medicine use, premature discontinuation and financial discontinuation (per-rupee OR ~ 1.00). Discontinuation sub-questions were answered only by those who had discontinued a course. \* $p<0.001$ , $p<0.01$ , \* $p<0.05$ .

Harmful practices were common: 69.0% reported sharing medicines, 58.5% using leftover medicine, and 53.2% discontinuing a course early; among those discontinuing, 80.6% cited perceived intolerance and 10.4% financial inability. Associations separated into two patterns (table 3). First, an apparently involuntary, poverty-related pattern: Baiga identity was associated with premature discontinuation (4.77, 3.39-6.70), stopping for perceived intolerance (4.43, 2.50-7.83), and financial discontinuation (6.81, 3.51-13.25; AUC 0.837), and lower per capita income independently predicted these outcomes. Notably, Baiga and lower-education respondents were less likely to stop because they perceived the illness had resolved (Baiga 0.12, 0.05-0.32; primary-or-less 0.09), suggesting non-completion was not driven by feeling better. Second, a knowledge-and-behaviour pattern: leftover-medicine use was associated with Baiga identity (1.93), lower income, and younger age, and medicine-sharing with younger age. Chronic disease was associated with use of human medicines in livestock (2.02) and with stopping when symptoms progressed (3.29). Demand-side behaviours (asking for antimicrobials, use without prescription, current household use) had no FDR-significant predictors.

### Actual treatment-seeking and cost: 30-day recall sub-study

In the sub-study (n=284; single-provider models n=280; government reference), Baiga identity was strongly associated with non-government care during actual illness (table 4). Relative to government facilities, Baiga individuals had higher odds of using JSS (16.94, 3.70-77.60) and informal practitioners (11.89, 2.23-63.53); the private contrast was elevated but not significant (5.88). The predicted probability of using a government facility was 1.6% for Baiga versus 18.1% for non-Baiga individuals, with JSS accounting for 70.7% of Baiga episodes (table 4). The small number of Baiga government users widened these intervals, so estimates are imprecise. Out-of-pocket cost for the episode (Z) exceeded usual monthly health expenditure (Q; paired Wilcoxon p<0.0001); median episode costs were modest at government and JSS providers but the upper tail was much higher at private and informal providers (up to INR 150 000; table 4; figure 1).

**Table 4:** 30-day treatment-recall sub-study.

(a) Multinomial regression of actual treatment location (reference: government)
| Predictor | OR (95% CI) | p (FDR) | Sig |
| --- | --- | --- | --- |
| **JSS vs government** |  |  |  |
| Baiga (ref non-Baiga) | 16.94 (3.70-77.60) | 0.009 | ** |
| Occupation: housewife (ref daily labour) | 8.88 (2.47-31.94) | 0.014 | * |
| **Informal practitioner vs government** |  |  |  |
| Baiga (ref non-Baiga) | 11.89 (2.23-63.53) | 0.042 | * |
| **Private clinic vs government** |  |  |  |
| Baiga (ref non-Baiga) | 5.88 (1.16-29.85) | 0.180 |  |

(b) Predicted probability of actual treatment location (%)
| Group | Government | JSS | Private | Informal |
| --- | --- | --- | --- | --- |
| Non-Baiga (ref) | 18.1 | 45.9 | 20.7 | 15.4 |
| Baiga | 1.6 | 70.7 | 11.0 | 16.6 |

(c) Monthly health expenditure (Q) vs treatment-episode cost (Z), median (IQR), INR
| Treatment location | n | Q monthly health expenditure | Z episode cost |
| --- | --- | --- | --- |
| Government | 53 | 100 (100-150) | 150 (100-200) |
| JSS | 134 | 100 (50-200) | 150 (50-400) |
| Private | 51 | 100 (100-150) | 200 (100-850) |
| Informal | 43 | 100 (100-200) | 200 (150-500) |

Paired Wilcoxon signed-rank test (Z vs Q, n=283): $p < 0.0001$ . Medians and IQRs given marked right-skew.

**Figure 1:**
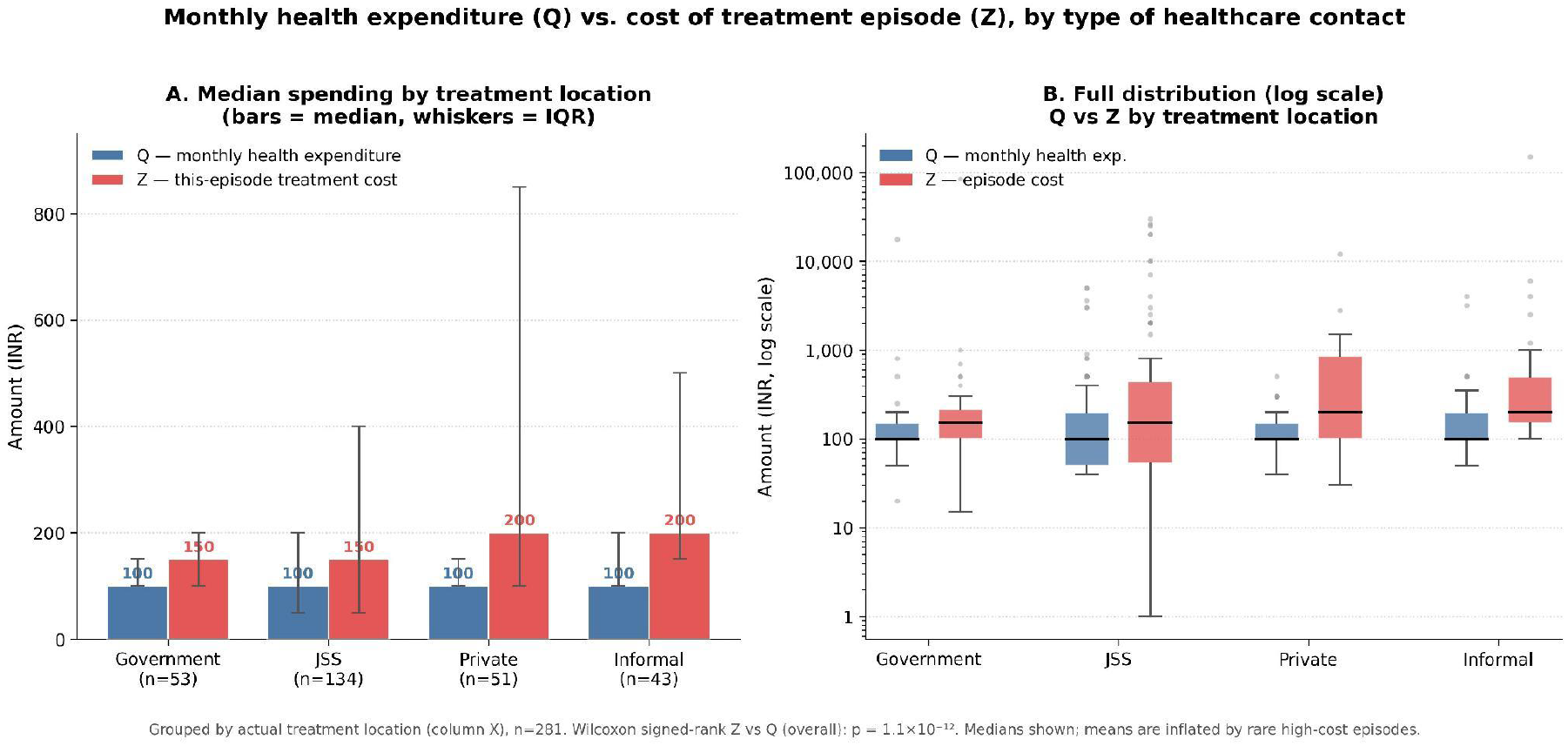
Monthly health expenditure (Q) and treatment-episode cost (Z) by treatment location. *Left: median Q and Z by provider. Right: distribution of episode cost (Z); the upper tail is far higher for private and informal providers. 30-day recall sub-study*.

### Sensitivity analysis

Refitting the sparse and separated models with Firth penalised regression reproduced the findings (appendix table A1). The discontinuation associations were essentially unchanged (Baiga: intolerance 4.28, financial 6.45, perceived resolution 0.13). In the sub-study, the large maximum-likelihood odds ratios attenuated but remained significant (Baiga vs government: JSS 11.31, informal 5.82, private 5.12; all p<=0.01).

## Discussion

In this population-level survey of a marginalised tribal community in central India, community identity - rather than income or education alone - was the most consistent correlate of where people sought care and of poverty-related antibiotic discontinuation. Three points follow.

First, government services appeared to reach Baiga residents poorly: Baiga identity was the main correlate of non-government care, and predicted government use during actual illness was very low (1.6%). Because the models adjusted for income and distance, this gap is unlikely to be explained by those factors alone and may reflect access and acceptability barriers. Residents who did not use government care relied on private or informal providers, which carried a higher risk of large out-of-pocket costs.

Second, the data suggest antibiotic non-completion in this setting is not a single behavioural phenomenon. A knowledge-and-behaviour pattern (medicine sharing, leftover use) coexisted with an apparently involuntary, poverty-related discontinuation pattern concentrated among Baiga and lower-income respondents, who were also less likely to stop because they felt better. With awareness of antimicrobials at 7.6%, interventions framed around “misuse” and awareness may have limited traction where non-completion is constrained by cost and access. These patterns support evaluating affordability-oriented, differentiated approaches - including support for course completion - rather than awareness messaging alone.

Third, use of the non-profit provider (JSS) did not differ between Baiga and non-Baiga residents, in contrast to government facilities, and JSS use increased with distance. This is consistent with more equitable reach by a trusted, community-based provider. However, this comparison must be read cautiously: the survey was conducted within JSS’s own catchment and recruited through JSS community health workers, which favours JSS use and limits inference about transferability. The finding is best regarded as hypothesis-generating, warranting evaluation in settings not anchored to a single provider.

### Limitations

This study has several limitations. The cross-sectional design precludes causal inference, and income and behavioural data are self-reported, with potential recall and social-desirability bias. Because participants were recruited on a catchment basis through JSS rather than by probability sampling, the sample may not represent all Baiga or Scheduled Tribe communities and is subject to selection effects related to engagement with JSS; in particular, the more equitable reach observed for JSS should be read in light of recruitment through its own community health network, and most villages contributed few respondents, so residual within-village correlation is expected to be small. AUC values for several models were modest (0.57-0.64), indicating unmeasured determinants (provider availability, supply-side practices, illness severity). The recall sub-study was small (n=284), with few Baiga using government facilities, producing wide intervals and separation that required Firth penalisation; episode cost is endogenous to provider choice. Provider-choice models excluded mixed-provider contacts, which may understate hybrid care-seeking.

## Conclusion

In this marginalised tribal population, antibiotic non-completion was associated with poverty and constrained access rather than only with awareness, and a community-based non-profit provider appeared to reach groups more equitably than government services. These associational findings support prospective evaluation of affordability-oriented, differentiated stewardship and of community-based delivery models, in studies designed to overcome the sampling limitations of the present work.

## Data Availability

Data are not publicly available owing to the risk of re-identifying members of a small Indigenous community. De-identified data may be available from the corresponding author on reasonable request, subject to approval by the Shaheed Hospital institutional ethics committee.

## Contributors

Conceptualisation: DK, KW, BC, RK. Methodology: DK, TS, KW, BC, RK. Formal analysis: RRN, DK. Data collection: GP, DS, IP. Writing (original draft): DK. Writing (review and editing): all authors. DK is the guarantor and had full access to the data and final responsibility for the decision to submit.

## Acknowledgements

We thank the communities of the Shivtarai, Semaria, and Bamhni areas, and the JSS field investigators and community health workers who facilitated data collection.

## Appendix

**Appendix table A1.** Maximum-likelihood (ML) versus Firth penalised-likelihood estimates for models with low events-per-variable or separation.

| Model | Predictor | ML OR | Firth OR (95% CI) | Firth p |
| --- | --- | --- | --- | --- |
| **Antimicrobial-discontinuation outcomes (n~640; 8 predictors)** |  |  |  |  |
| O8 Stopping, perceived intolerance | Baiga | 4.43 | 4.28 (2.44-7.51) | <0.001 |
|  | Chronic disease | - | 2.31 (1.06-5.07) | 0.007 |
| O9 Discontinuation, financial inability | Baiga | 6.81 | 6.45 (3.38-12.29) | <0.001 |
|  | Per capita income (per 1000) | - | 0.88 (0.83-0.94) | <0.001 |
| O10 Stopping, perceived resolution | Baiga | 0.12 | 0.13 (0.05-0.33) | <0.001 |
|  | Edu: primary or less | 0.09 | 0.09 (0.03-0.26) | <0.001 |
|  | Edu: high school | 0.15 | 0.16 (0.06-0.44) | <0.001 |
|  | Chronic disease | 3.29 | 3.22 (1.72-6.03) | <0.001 |
| **30-day recall sub-study: actual treatment location (reference government)** |  |  |  |  |
| JSS vs government | Baiga | 16.94 | 11.31 (2.91-44.02) | <0.001 |
| Informal vs government | Baiga | 11.89 | 5.82 (1.23-27.45) | 0.008 |
| Private vs government | Baiga | 5.88 | 5.12 (1.13-23.32) | 0.008 |

ML=maximum likelihood. OR=odds ratio. Firth p-values are penalised likelihood-ratio tests. For the discontinuation models, ML point estimates are shown but their CIs were unstable (a singular information matrix arose for the financial-inability model); "-" denotes a predictor outside the FDR-significant set. Sub-study ML estimates are from the four-way multinomial model; Firth models were fitted as one-versus-reference binary models with a parsimonious predictor set.

